# Hi-C detects genomic structural variants in peripheral blood of pediatric leukemia patients

**DOI:** 10.1101/2021.10.01.21264442

**Authors:** Claire Mallard, Michael J Johnston, Anna Bobyn, Ana Nikolic, Bob Argiropoulos, Jennifer A Chan, Gregory MT Guilcher, Marco Gallo

**Affiliations:** Arnie Charbonneau Cancer Institute, Cumming School of Medicine, University of Calgary, Calgary, AB; Alberta Children’s Hospital Research Institute, Cumming School of Medicine, University of Calgary, Calgary, AB; Department of Molecular Biology and Biochemistry, Cumming School of Medicine, University of Calgary, Calgary, AB; Department of Medical Genetics, Cumming School of Medicine, University of Calgary, Calgary, AB; Department of Oncology, Cumming School of Medicine, University of Calgary, Calgary, AB; Department of Pediatrics, Cumming School of Medicine, University of Calgary, Calgary, AB

## Abstract

B-cell acute lymphoblastic leukemia (B-ALL) is often driven by chromosome translocations that result in recurrent and well-studied gene fusions. Currently, fluorescent in-situ hybridization probes are employed to detect candidate translocations in bone marrow samples from B-ALL patients. Recently Hi-C, a sequencing-based technique originally designed to reconstruct the three-dimensional architecture of the nuclear genome, was shown to effectively recognize structural variants. Here, we demonstrate that Hi-C can be used as a genome-wide assay to detect translocations and other structural variants of potential clinical interest. Structural variants were identified in both bone marrow and peripheral blood samples, including an *ETV6-RUNX1* translocation present in one pediatric B-ALL patient. Our report provides proof-of-principle that Hi-C could be an effective strategy to globally detect driver structural variants in B-ALL peripheral blood specimens, reducing the need for invasive bone marrow biopsies and candidate-based clinical tests.

## INTRODUCTION

B-cell acute lymphoblastic leukemia (B-ALL) is a malignancy of the bone marrow and blood that results in uncontrolled proliferation of B-cell lineage progenitors. The frequency distribution of B-ALL diagnoses is bimodal, with peaks in childhood and in adults around 50 years of age (reviewed in^1^). New treatment approaches have considerably improved overall outcomes in children, with 98% of pediatric patients going into remission following treatment and a cure rate of 90%. On the other hand, clinical management of adult B-ALL has proven more challenging, with median overall survival of 11 months.

At the genomic level, B-ALL is primarily driven by translocations and other structural variants (SVs) that are thought to originate in B-lymphocyte progenitor cells. Some recurrent translocations occur with high frequency in the patient population and can result in tumorigenic, gain-of-function fusion proteins. These genomic events include t(9;22) (a translocation between chromosome 9 and chromosome 22, also known as the Philadelphia chromosome) resulting in *BCR-ABL1* fusion; t(12;21), resulting in the *ETV6-RUNX1* fusion; t(1;19), resulting in the *TCF3-PBX1* fusion; and rearrangements of the *MLL* gene with many fusion partners (reviewed in^2^). Detection of specific translocation(s) and SVs present at diagnosis allows molecular subtyping of the B-ALL case, which informs patient prognosis and guides therapy selection.

Clinical tests for the detection of translocations mostly rely on fluorescent in-situ hybridization (FISH) of bone marrow samples. Bone marrow cells are obtained via pelvic bone tap, which is an invasive and painful procedure. Since this is a candidate-based clinical test, multiple FISH probes must be used to assay for candidate translocations. Karyotyping is also performed on these samples, but this method may miss SVs that are not big enough to be detected. Being able to capture different types of SVs – including smaller events – in one individual test could improve speed and precision of diagnosis.

In this report, we test the potential of Hi-C^3^ – a chromosome conformation capture technique – to provide effective genome-wide information on the SV status of B-ALL samples. Hi-C was originally designed to assess physical interactions between non-contiguous chromosome regions^4,5^. This information is then used to computationally reconstruct three-dimensional (3D) genome architecture (i.e. how DNA folds within the nucleus). In essence, Hi-C involves crosslinking DNA within cells to create a snapshot of which genomic regions physically interact with each other. When libraries of interacting regions are sequenced very deeply, it is possible to reconstruct both large-scale genomic structures (domains and compartments) and finer features like chromatin loops formed by cis regulatory regions interacting with their target promoters^4,6^.

Hi-C can visualize major SVs in a sample^7^. For instance, translocations would appear as interactions between regions normally situated on different chromosomes with a characteristic butterfly appearance on a Hi-C contact map (**Fig 1**). Recent publications have shown that Hi-C enables detection of SVs even with relatively shallow sequencing of the libraries^7,8^.

**Fig 1.**
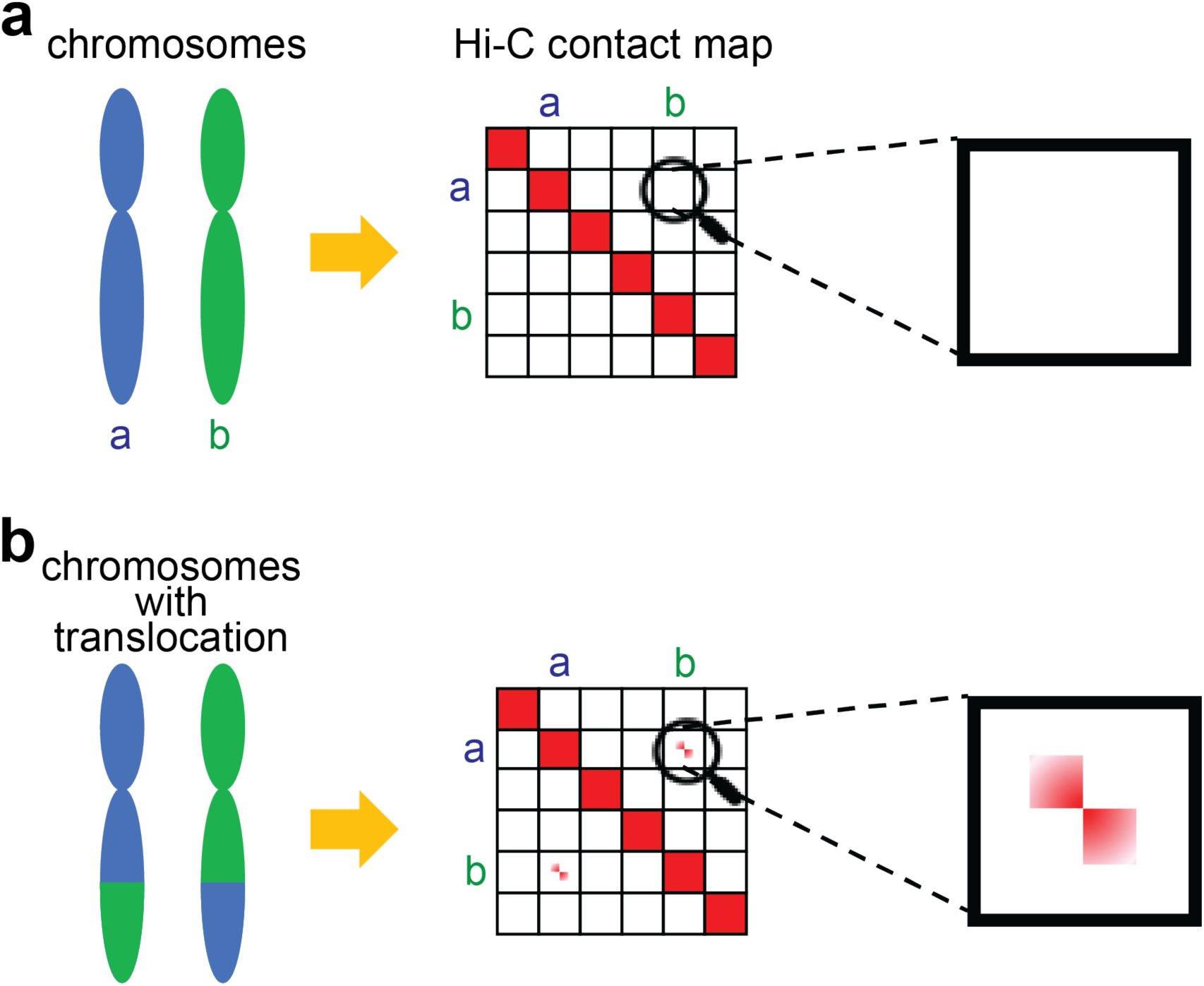
Identification of chromosomal translocations with Hi-C. (a) Diagram illustrating the expected Hi-C contact frequency maps in the case of a genome with no translocations. A normal karyotype will result in minimal interchromosomal contacts. (b) Diagram illustrating the expected Hi-C contact frequency maps in the case of a genome a balanced translocation. Elevated interchromosomal contacts would be observed. A balanced translocation would appear as a contact with a characteristic “butterfly” appearance (enlarged inset).

Here, we assess the feasibility of using shallow (low-coverage) Hi-C to infer translocations and other SVs in a discovery cohort of children with B-ALL. We also test the feasibility of detecting SVs by performing Hi-C on peripheral blood of B-ALL patients as a less-invasive alternative to using bone marrow aspirates.

## RESULTS

### Hi-C datasets of bone marrow and peripheral blood samples from pediatric B-ALL patients

We generated Hi-C libraries for 5 samples from 4 pediatric B-ALL patients (**Table 1**) with an average contact resolution of ∼35 kb. For all the relevant statistics related to the Hi-C libraries, please see **Supplemental Table S1**. For patient 4437, we generated Hi-C data on both bone marrow cells (4437M) and peripheral blood cells (4437B) to test our hypothesis that translocations and structural variants can be detected in leukemic blasts that circulate in the peripheral blood. For all the other patients, we generated Hi-C datasets from their peripheral blood. For each patient, we also had clinical data about their B-ALL diagnosis using FISH (**Table 2**). Our cohort included 2 males and 2 females. Samples from 3 patients were collected at initial diagnosis of leukemia. The sample from patient 4441 was obtained when that individual was in remission.

Hi-C maps generated from all samples were informative of the overall genomic structure of all samples (**Fig 2a-e**). They all show strong interactions primarily within each chromosome, as expected, including stronger interactions that preferentially occur within the p-arm and q-arm of each chromosome. However, it is also possible to notice putative interactions between chromosomes, which likely represent SVs. A candidate SV, for instance, appears as a translocation between the q-arm of chromosome 1 and chromosome 9 in patient 4440 (**Fig 2d**).

**Table 1:**
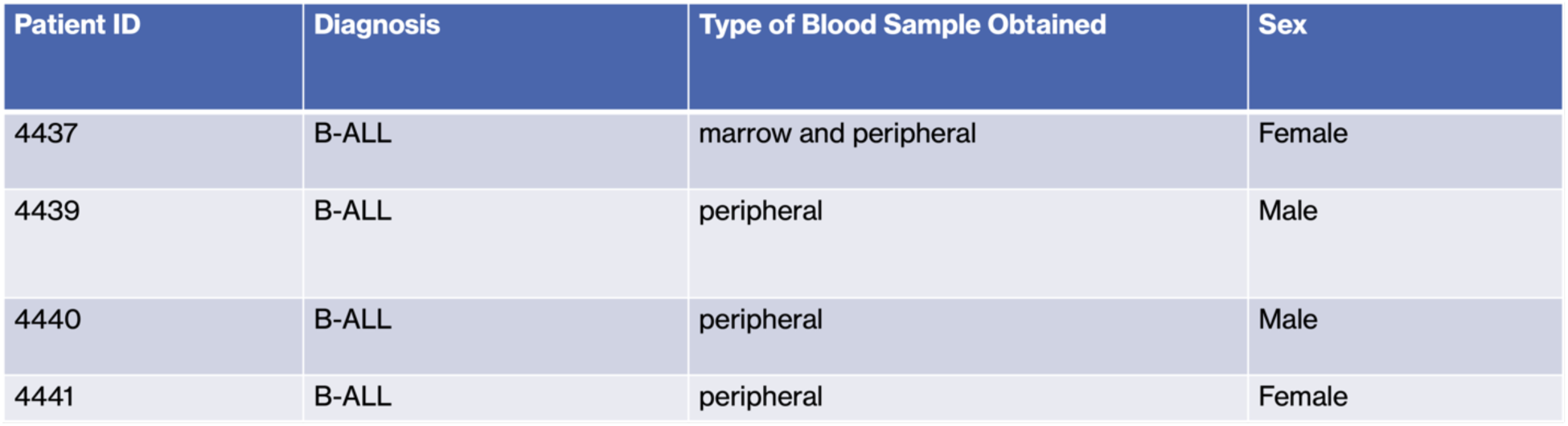
Patient metadata.

| Patient ID | Diagnosis | Type of Blood Sample Obtained | Sex |
| --- | --- | --- | --- |
| 4437 | B-ALL | marrow and peripheral | Female |
| 4439 | B-ALL | peripheral | Male |
| 4440 | B-ALL | peripheral | Male |
| 4441 | B-ALL | peripheral | Female |

**Table 2.**
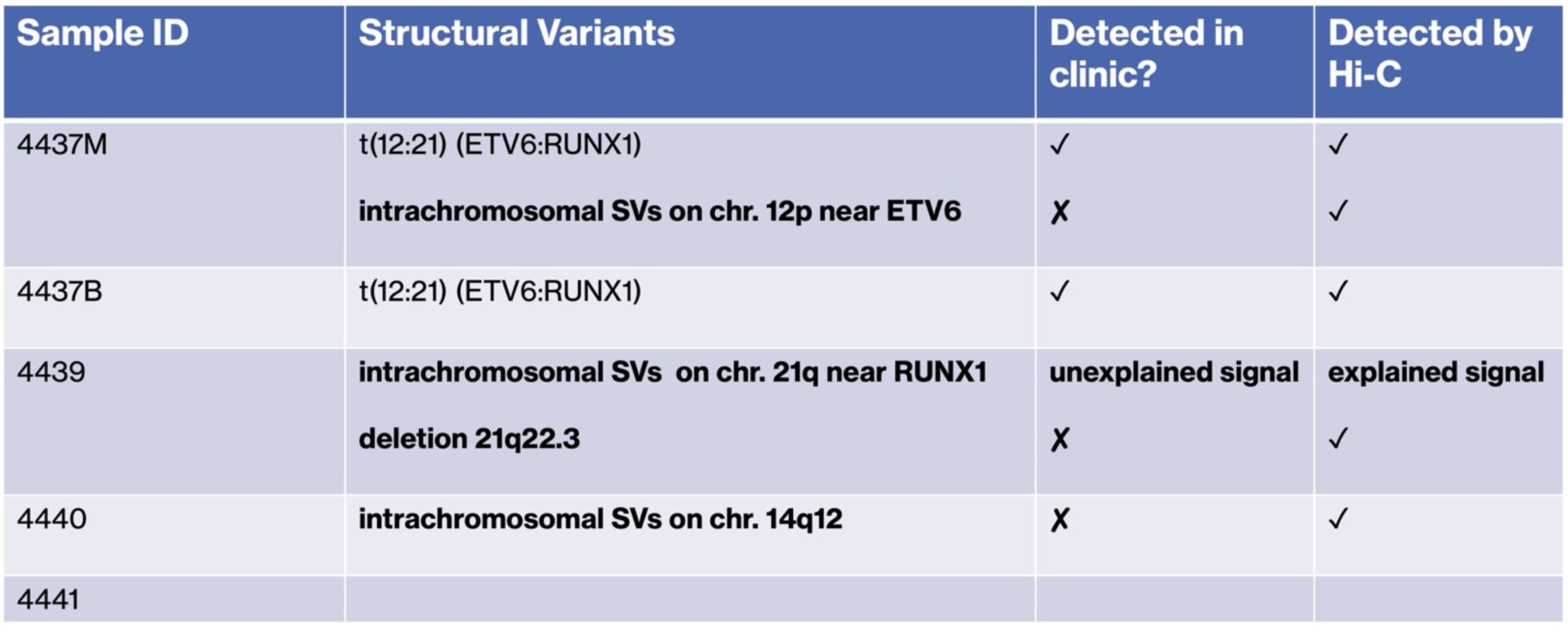
Comparison of structural variants detected by conventional cytogenetic analysis and Hi-C.

**Fig 2.**
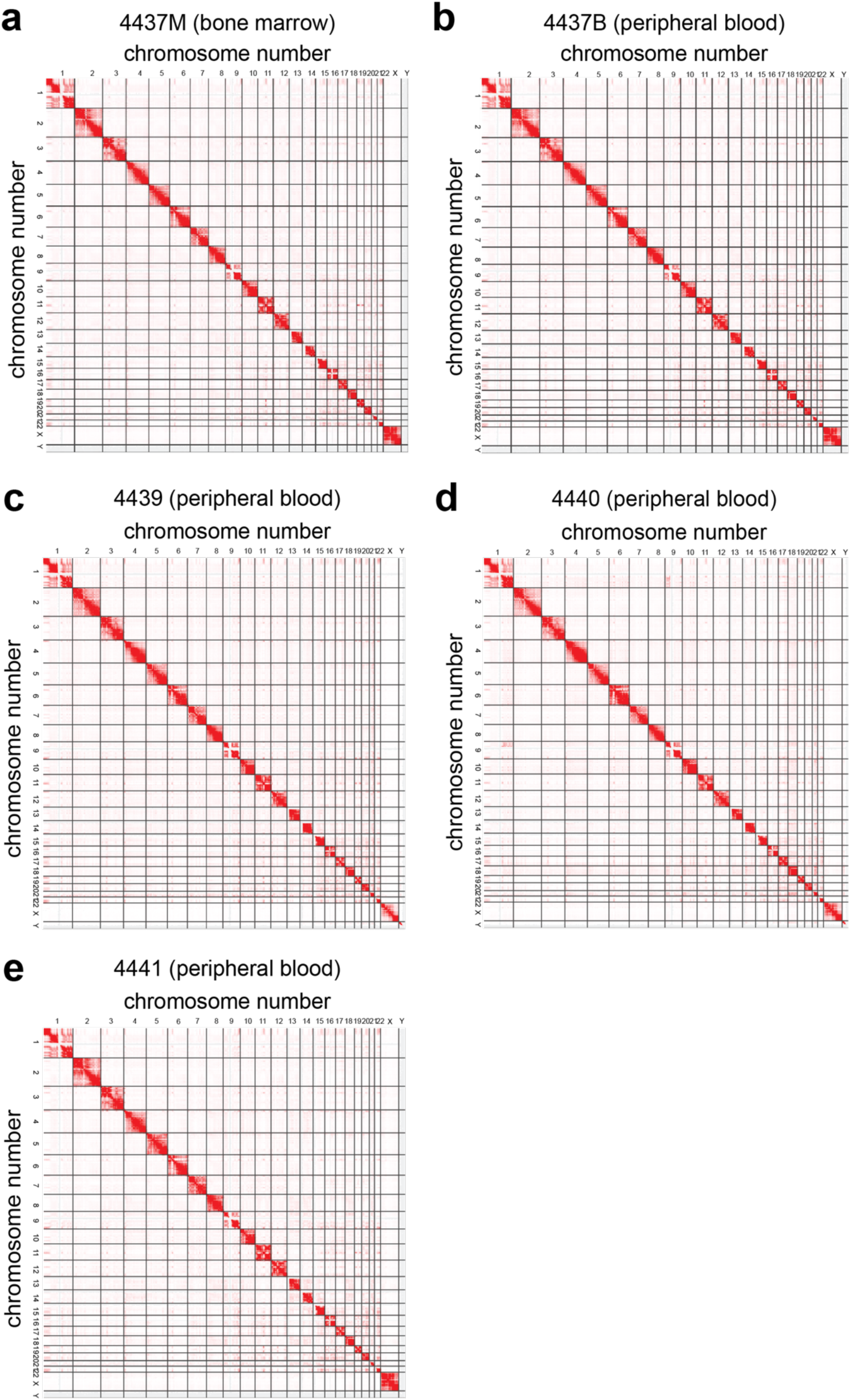
Hi-C contact maps for 5 samples collected from pediatric B-ALL patients. (a) Contact map for Hi-C data generated from the bone marrow sample of patient 4437. (b) Contact map for Hi-C data generated from the peripheral blood sample of patient 4437. (c) Contact map for Hi-C data generated from the peripheral blood sample of patient 4439. (d) Contact map for Hi-C data generated from the peripheral blood sample of patient 4440. (e) Contact map for Hi-C data generated from the peripheral blood sample of patient 4441.

We uploaded the processed Hi-C maps for all 5 samples to the WashU Epigenome Browser (https://wangftp.wustl.edu/hubs/gallo_B-ALL) to facilitate public access to our datasets and their downstream analyses by users. The raw Hi-C data are also available (see Data Availability section). We hope these datasets will be a resource for the community.

### Identification of translocations using Hi-C datasets

As a first test of our datasets, we wanted to determine if B-ALL related translocations could be detected using Hi-C data from the bone marrow sample we profiled. Clinical FISH probes confirmed that leukemic blasts from patient 4437 were positive for the *ETV6-RUNX1* translocation, which is a driver mutation for B-ALL and occurs via a balanced exchange of DNA between chromosome 12 and 21. Focusing on these chromosomes, we were able to identify this translocation, which appeared as a butterfly shaped signal, as expected (**Fig 3a**). Furthermore, our datasets allowed identification of the precise breakpoints of the *ETV6-RUNX1* translocation in this patient. The Hi-C contact matrix showed breakpoints corresponding to intron 5 of *ETV6* and intron 2 of *RUNX1* (**Fig 3b**). These results support previous findings on this well-researched translocation.

**Fig 3.**
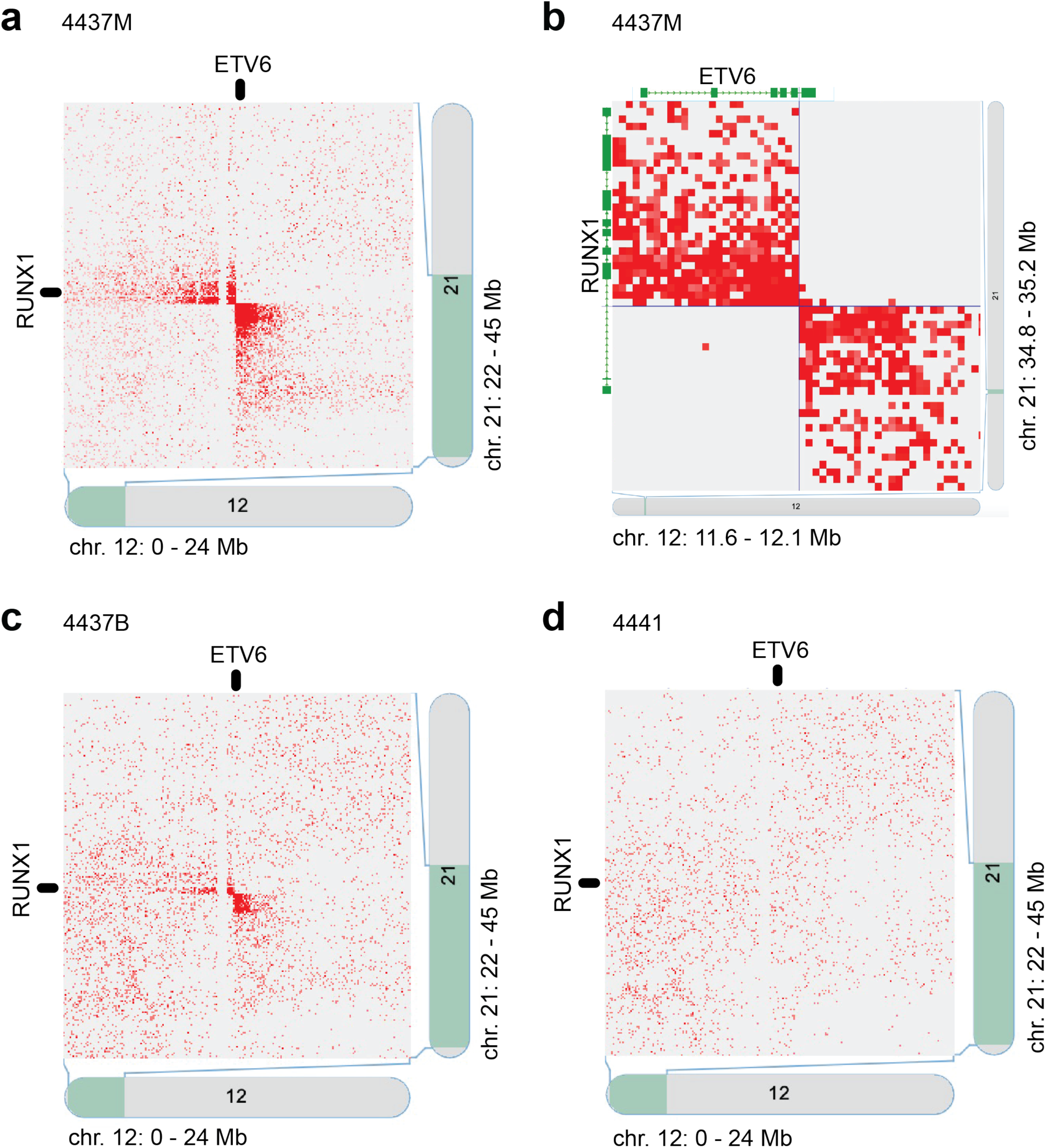
The ETV6:RUNX1 translocation detected in bone marrow and peripheral blood Hi-C contact maps. (a) The interchromosomal contact map showing interactions between chr. 12 (0 – 24 Mb) and chr. 21 (22 – 45 Mb) for the bone marrow sample of patient 4437. The bright red butterfly shape shows the balanced translocation at the ETV6 locus (chromosome 12) and the RUNX1 locus (chromosome 21) (b) Higher magnification view of the *RUNX1-ETV6* translocation detected by Hi-C in the bone marrow sample of patient 4437 (previous panel). The *ETV6* and *RUNX1* genes are shown to illustrate how the data can clearly identify the breakpoint of the translocation. The breakpoint for *ETV6* occurs in intron 5 and the breakpoint for *RUNX1* occurs in intron 2. (c) Interchromosomal contact map showing interactions between chromosome 12 (0 – 24 Mb) and chromosome 21 (22 – 45 Mb) for the peripheral blood sample of patient 4437. The bright red butterfly shape shows the balanced translocation at the ETV6 locus (chromosome 12) and the RUNX1 locus (chromosome 21). (d) Interchromosomal contact map for chromosome 12 (0 – 24 Mb) and chromosome 21 (22 – 45 Mb) for the peripheral blood sample of patient 4441. The absence of abnormal signal signifies that no translocation between these loci is present in this sample.

Next, for the same patient, we looked for that same translocation using Hi-C data from peripheral blood. Although peripheral blood has lower density of leukemic blasts than the bone marrow, we were still able to identify a significant signal at the expected site of translocation (**Fig 3c**). This finding is further supported by comparing the signals observed in the translocation-positive patient (4437) and a translocation-negative patient (4441). At the ETV6-RUNX1 intersection locus for patient 4441, there is no significant signal from the peripheral blood Hi-C data (**Fig 3d**). Altogether, our results support the notion that shallow Hi-C has sufficient sensitivity to detect SVs, even in peripheral blood samples from B-ALL patients.

### Identification of complex SVs in pediatric B-ALL samples using Hi-C

Having established that shallow Hi-C enables the detection of known B-ALL translocations, we then wondered if we could use our datasets to identify other SVs in the genomes of our patients. Examining our datasets, we were able to identify SVs that were not reported following clinical tests for the same patients (**Table 2**). These SVs appeared as strong signals off the diagonal of Hi-C contact matrices. Among the signals identified were candidate tandem duplications, inversions and deletions, but further analyses would be required to fully characterize each SV. However, the detection of abnormalities and the genes involved at these loci are a starting point for further analysis into these cancer genotypes that would not be possible to explore using only clinical FISH probes.

For patient 4439, complex intrachromosomal structural variants were detected around the *RUNX1* locus on chromosome 21 (**Fig 4a**). Some cytogenic abnormalities were detected near this locus in clinical analysis, but Hi-C was able to specifically identify the breakpoints. It is possible that this SV could affect *RUNX1* expression, but we cannot confirm this hypothesis with the data we currently have. Further, the effects seen line up closely with a known B-ALL cytogenetic subgroup known as iAMP21 - intrachromosomal amplification of chromosome 21^9^. Further, this subgroup has a 3-fold higher relapse rate and a lower 5-year survival (71%) than other B-ALL patients^9^, making the rapid diagnosis and characterization of the leukemia critical for proper diagnosis and clinical management of these high-risk patients.

**Fig 4.**
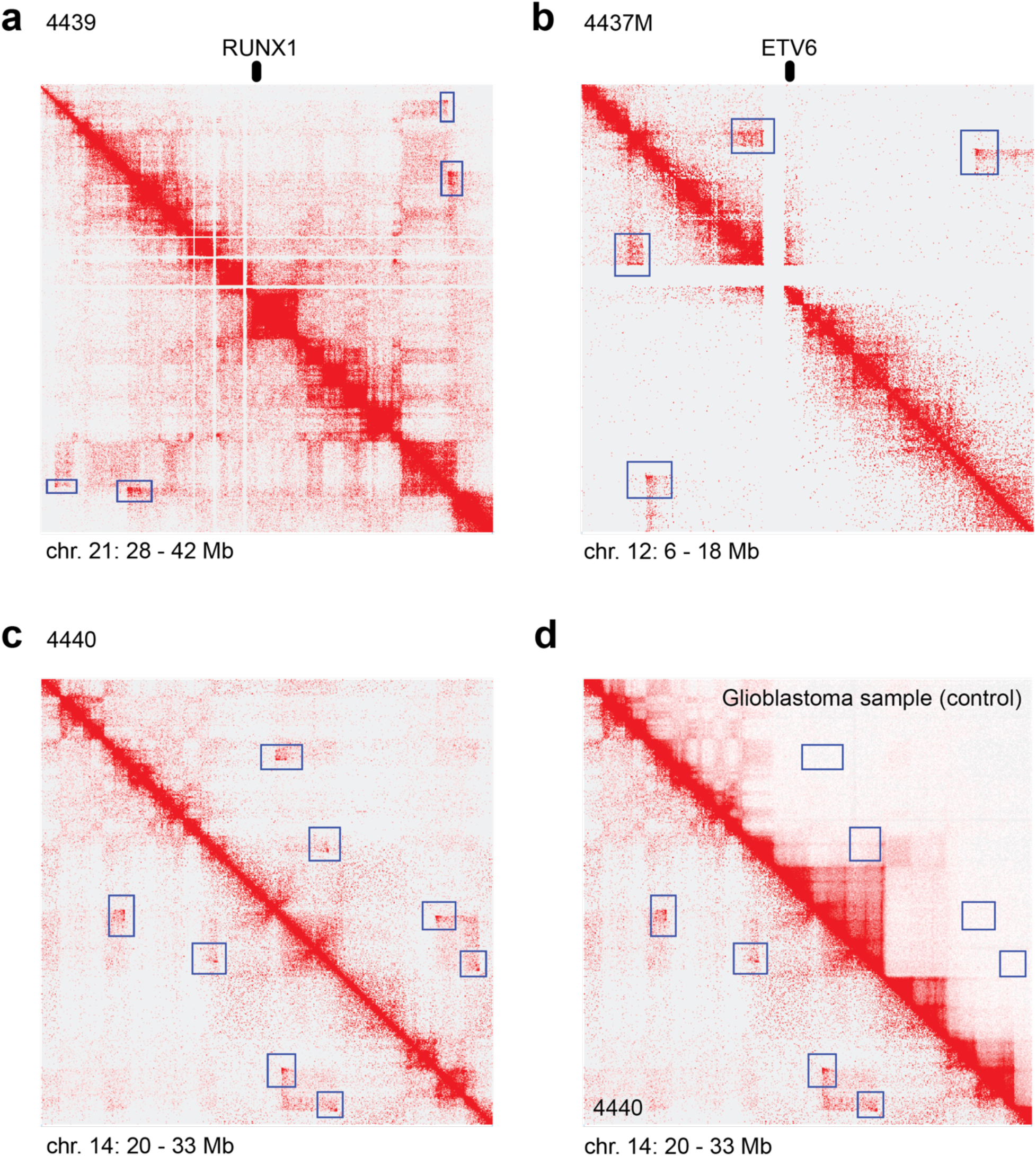
Hi-C enables genome-wide identification of structural variants. (a) Intrachromosomal structural variants near the *RUNX1* locus on chromosome 21 (28 – 42 MB) for patient 4439. (b) Intrachromosomal structural variants near the *ETV6* locus on chromosome 12 (6 – 18 MB) in the bone marrow sample of patient 4437. (c) Intrachromosomal structural variants on chromosome 14 (20 – 33 MB) for patient 4440. (d) Intrachromosomal structural variants on chromosome 14 (20 – 33 MB) for patient 4440 (bottom-left triangle) compared with the same region on a GBM patient Hi-C contact map (top-right triangle). For all panels, the putative structural variants are highlighted in blue boxes.

Further structural variants were detected in the bone marrow Hi-C contact matrix for sample 4437M, in which we previously detected the *ETV6-RUNX1* translocation (**Fig 4b**). These abnormalities were detected as discontinuous, long-range intrachromosomal interactions along chromosome 12 near the *ETV6* locus, suggesting that the local chromatin environment around ETV6 may be further disrupted independent of the ETV6-RUNX1 fusion. Further, the breakpoints of another potential SV (lower interaction box in **Fig 4b**) correspond to the genes *APOBEC1* and *LMO3*. Both genes have been identified as important for leukemias in previous studies. *APOBEC1* encodes a cytosine deaminase and has been linked to cancer development in acute myeloid leukemia^10^. *LMO3* codes for a transcription factor and oncogene that contributes to the etiology of T-cell acute lymphoblastic leukemia^11^.

We also identified a complex SV on the p-arm of chromosome 14 of the sample from patient 4440 (**Fig 4c**). This cluster of abnormal Hi-C signals is consistent with a chromothripsis event, in which the chromosome was partially shattered, and many gene loci were altered all at once. This SV was not observed in previously published Hi-C data from a glioblastoma sample (G523)^12^, which has non-altered chromosomal structure in this region (upper half of **Fig 4d**).

### 3D modeling of B-ALL chromatin interactions

In addition to large-scale SVs, Hi-C provides information on chromatin interactions along chromosome regions that do not harbor obvious SVs^4,5^. 3D chromatin interactions result in the formation of domains and loops, which are important factors for gene expression and determination of cell states^13^. We decided to test whether our shallow Hi-C data were sufficient to probe chromatin interactions in B-ALL patient samples. To achieve this goal, we used CSynth^14^, a computational tool that generates 3D models of chromosomes based on data generated from chromosomal conformation techniques like Hi-C. We modelled a region of chromosome 12 (77 – 86 MB) and looked for differences in loop structure between bone marrow cells (**Fig 5a**) and peripheral blood cells (**Fig 5b**) for patient 4437. We expected to see differing loop structure between the two, since 3D chromatin looping structures are cell-type specific. We highlighted two pairs of loop anchors corresponding to two loops: 77.1 – 77.8 MB and 78.1 – 79.4 MB (**Fig 5a,b**; green and red, respectively). In Hi-C, the intensity of the loops was higher in the bone marrow data, likely due to the different density of bone-marrow specific cells, such as B-lymphocytes. This was supported by the 3D models which showed tighter and more distinct loops in the bone marrow model than in the peripheral blood model, as seen by the closer proximity of the green and red loop boundaries.

**Fig 5.**
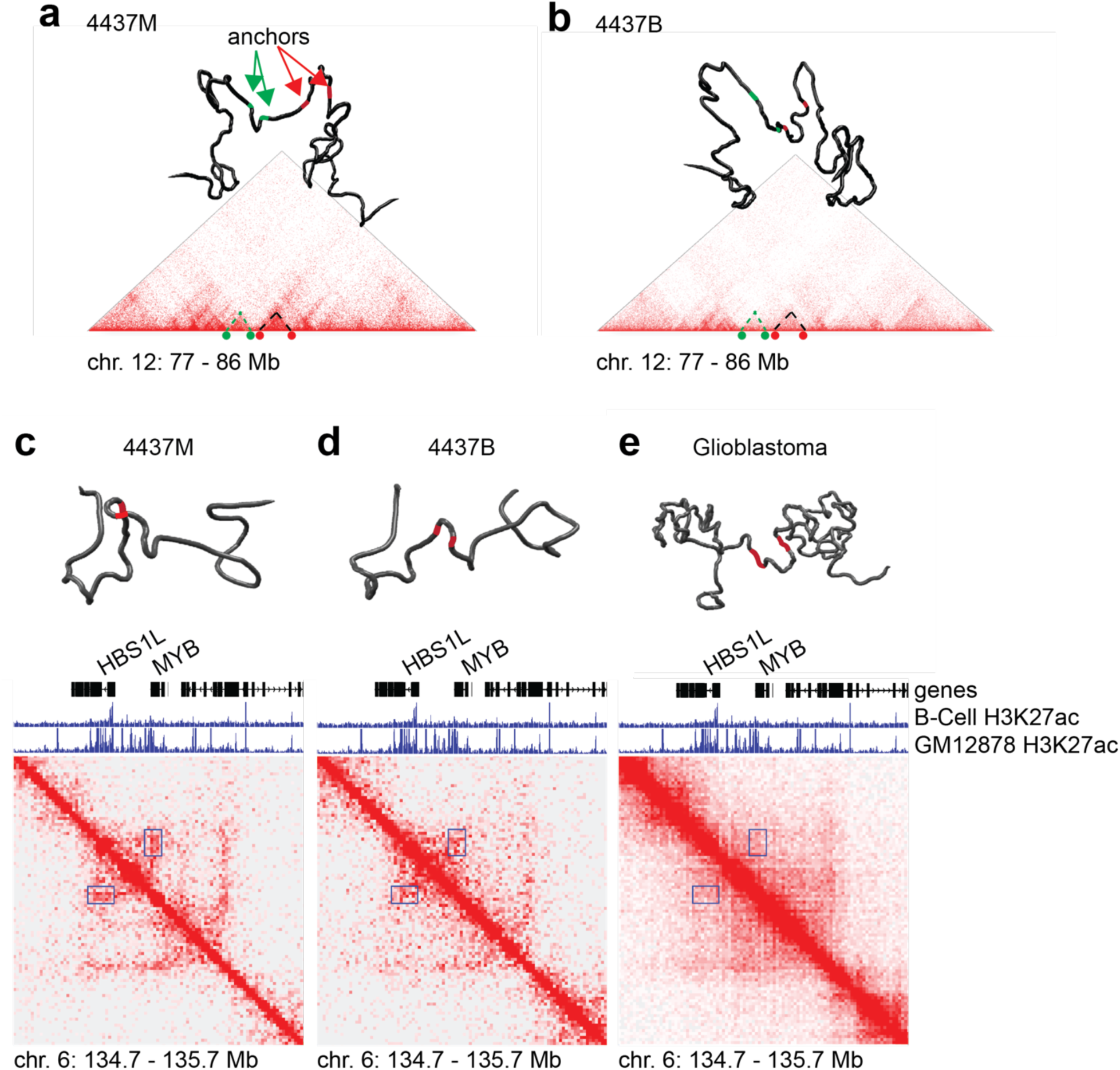
3D modeling of shallow Hi-C data reveals information on gene regulation. (a-b) Hi-C based 3D models of chromosome 12 (77 – 86 MB) from patient 4437 displaying both bone marrow (a) or peripheral blood (b). Green highlights the loop boundaries for the loop spanning 77.1 – 77.8 MB and red highlights the loop boundaries for the loop spanning 78.1 – 79.4 MB. (c-e) Hi-C contact matrices and 3D models of chromosome 6: 134.7 – 135.7 MB for patient 4437 bone marrow (c) patient 4437 peripheral blood (d) and a control glioblastoma patient (e). The loop formed between an H3K27ac enhancer region and the gene MYB is highlighted in the blue box on the Hi-C matrix and by red loop boundaries in the 3D model. ChIP-seq tracks for the histone mark H3K27ac for a primary B cell sample and a cell line (GM12878) are shown in each panel.

Next, we wanted to see if this phenomenon would be apparent for specific enhancer to gene loops related to leukemias. We identified putative cis regulatory regions using chromatin immunoprecipitation with sequencing (ChIP-seq) data for B cells and for GM12878, a lymphoblastoid cell line, generated by ENCODE^15^. Specifically, we looked at acetylation of histone 3 on lysine 27 (H3K27ac), which marks enhancer and promoter regions. We found a strong loop present from the bone marrow Hi-C between a putative enhancer marked by H3K27ac and the proto-oncogene *MYB* which is over-expressed in leukemias^16^. In the Hi-C contact matrix for patient 4437 bone marrow, there was a strong signal at the interaction locus for the enhancer and *MYB* (**Fig 5c**). This corresponded to an obvious loop structure in the 3D model (**Fig 5c**). In the peripheral blood Hi-C contact matrix for the same patient, there was a signal present, but with lower intensity. The 3D model also showed the loop, but the boundaries were farther apart (**Fig 5d**). Finally, this loop was not present in the Hi-C contact matrix of a previously published glioblastoma sample (**Fig 5e**). These results demonstrate that shallow Hi-C datasets of clinical samples are sufficient to reconstruct the 3D milieu of specific genomic regions that are relevant to disease etiology.

## DISCUSSION

Hi-C has been shown to be an effective genomic approach to solve the 3D architecture of the genome, and also to identify large SVs^7,8^. We reasoned that the unbiased, genome-wide assessment of structural variation offered by Hi-C could be an asset in the diagnosis and molecular subtyping of malignancies driven by defined SVs, especially translocations. We therefore decided to focus our efforts on B-ALL, which is very often driven by translocation events that generate gene fusions that maintain malignant cells in progenitor states.

Balanced translocations can easily be visualized in Hi-C contact maps by virtue of their characteristic “butterfly” shape. One of our patients had a canonical translocation between chromosomes 12 and 21, resulting the in the *ETV6-RUNX1* gene fusion. This translocation was confirmed by FISH. Our shallow Hi-C approach was able to clearly identify this translocation in the bone marrow sample by simple visual inspection. Importantly, we were also able to detect this translocation in the peripheral blood sample from the same patient. Additionally, we successfully identified other putative SVs in the peripheral blood samples of other 2 patients. The blood sample from an additional patient was collected when the child was in remission and, as expected, did not have obvious abnormal Hi-C signals. Overall, our data support the suitability of shallow Hi-C as a tool to detect known and unknown SVs in B-ALL genomes.

Although we only had one patient-matched pair of bone marrow and peripheral blood specimens, our results provide proof-of-principle that shallow Hi-C can be successfully applied to peripheral blood samples to detect recurrent translocations in B-ALL patients. The success of this approach depends upon on the burden of leukemic blasts in circulation and could correlate with disease stage. We expect that simple refinements of the approach we describe could significantly improve the power to detect translocations using peripheral blood. For instance, a simple solution would be to enrich for leukemic blasts by performing fluorescence-activated cell sorting directly on peripheral blood^17,18^. This approach would reduce the need for bone marrow biopsies, which can be painful procedures.

In patients who did not have the recurrent *ETV6-RUNX1* translocation, we also detected putative SVs near *ETV6* and *RUNX1*. This finding is interesting, because it suggests that there may be alternative mechanisms of disrupting proper expression of these genes in addition to the canonical translocation. Hi-C allows the visualization of large genomic abnormalities and could provide information of the potential disruption of transcriptional programs associated with these genes. Leukemias that converge on disruption of transcriptional pathways mediated by *ETV6* and *RUNX1*, whether through translocations or other means, could be identified.

In addition to providing unbiased, genome-wide contact data, Hi-C may also be more cost-effective than clinical FISH probes. To detect SVs, Hi-C can be performed at low resolution for a cost of $200-$1000 per sample (range depends on desired resolution and sequencing instruments available), offering cost effectiveness and improved sensitivity for unanticipated genomic events. Therefore shallow Hi-C is cost-competitive with other sequencing-based methods that are currently in use for diagnosis and molecular subtyping of leukemias in some hospitals^19^.

Shallow Hi-C does have some limitations. These include the specialized informatic analysis required and its inability to detect very small SVs (e.g. single-gene duplications or very small deletions). Hi-C output is also relatively qualitative, and visual inspection is required for inferences of genomic structural anomalies. However, future studies could lead to standard computational pipelines for accurate identification of SVs using Hi-C.

Additional studies with larger cohorts of patients will be required, but our results suggest that shallow Hi-C could be used to reinforce standard of care testing (flow cytometry and cytogenetics) and hopefully implemented in the clinic to test for translocations using peripheral blood of B-ALL samples. The standardization of Hi-C library preparation and its related computational pipelines, and wide availability of sequencing in clinical settings, make this option worth exploring.

## METHODS

### Patient samples

Collection of all samples was based on written informed consent and their use was approved by the Health Research Ethics Board of Alberta Cancer Committee (certificate number HREBA.CC-18-0169).

### Generation of Hi-C datasets

Peripheral or marrow blood samples were collected from patients with B-ALL at the Alberta Children’s Hospital. Blood samples were diluted 1:1 with PBS + 2% FBS. Peripheral blood mononuclear cells were isolated using SepMate columns (Stemcell Technologies). Cells were rinsed and resuspended to 25 mL in PBS + 2% FBS. Cell density was measured using a Countess cell counter (ThermoFisher). One to five million cells were then used as input for Hi-C library construction using the Arima Hi-C Kit (Arima Genomics), including KAPA Hyper Prep indexing and library amplification.

Samples were sequenced 2×150bp on a NextSeq 500 instrument (Illumina) at the Center for Health Genomics and Informatics (CHGI) at the University of Calgary. Reads were processed using Juicer^20^, aligning to reference genome hg38 and the restriction enzyme DpnII, to generate Hi-C contact matrices (.hic files). Contact matrices were visualised using Juicebox^21^.

### SV detection

We considered that genomic structural variants would appear as long-range interactions on a Hi-C contact map. Using Juicebox, we visually inspected the whole genome contact map using balanced normalization at 100-kbp resolution for abnormally high interchromosomal signals located >5 Mbp apart. 50-kbp resolution was used to inspect abnormal intrachromosomal signals. Additionally, putative SV signals were inspected to ensure the Observed/Expected ratios were much greater than 1. Finally, putative SV signals were compared to negative control Hi-C datasets in which leukemic disease had cleared (sample 4441) or which would not be expected to share leukemic genomic alterations (published glioblastoma dataset from sample G523^12^).

### 3D modeling of Hi-C data

Hi-C contact matrices and BED files containing loop loci of interest were loaded into CSynth^14^ (csynth.org). Default parameters were used, with the following exceptions: Push apart force: 3e-4; contact force: 52; diameter: 20.

### H3K27ac ChIP-seq datasets

We accessed H3K27ac fold enrichment traces from ENCODE^15^ through Juicebox with the following identifiers: ENCFF696PMM (Homo sapiens B cell female adult (27 years of age), web link: https://www.encodeproject.org/files/ENCFF696PMM/); ENCFF340JIF (Homo sapiens GM12878, web link https://www.encodeproject.org/files/ENCFF340JIF/).

## Data Availability

All Hi-C datasets generated for this study are available to the community. The processed Hi-C data can be loaded into the WashU Epigenome Browser from the following link: https://wangftp.wustl.edu/hubs/gallo_B-ALL. The raw Hi-C data have been uploaded at the EGA repository (accession number EGAS00001005605).

## AUTHOR CONTRIBUTIONS

MG and GMTG conceived the study. GMGT and JAC collected and banked the patient specimens. CM, MJJ, AB and BA performed the experimental activities. MG, CM and MJJ wrote the manuscript. All co-authors contributed to editing the manuscript.

## ACKNOWLEDGMENTS

This project was funded by a Young Investigator Award in Cell and Gene Therapy for Cancer grant from the Alliance for Cancer Gene Therapy (ACGT) to MG; a Discovery Grant from the Natural Science and Engineering Research Council (NSERC) to MG; a Canada Research Chair from the Government of Canada to MG; a USRA studentship from NSERC to CM; a postdoctoral fellowship from the Canadian Institutes of Health Research (CIHR) to MJJ; support from the Alberta Children’s Hospital Foundation and the Kids Cancer Care Foundation of Alberta to JAC for the Brain Tumor and Tissue Bank.

